# Automated Interpretation of EEG Reports Using a Large Language Model with Structured Confidence Outputs

**DOI:** 10.64898/2026.07.07.26357190

**Authors:** Wanying Tian, Steven Bergner, Alexander Moiseev, Fred Popowich, George Medvedev, Mark P. Richardson, Roman Rodionov, Pengcheng Xi, Sam M. Doesburg, Urs Ribary, Joel S. Winston, Vasily A. Vakorin

## Abstract

**Objective:** To evaluate a large language model (LLM) pipeline for extracting structured diagnostic labels and confidence levels from unstructured free-text EEG reports, addressing the barrier of narrative data analysis.

**Methods:** We developed a hierarchical schema classifying reports for four abnormality types using a four-point confidence scale. Two certified EEG technologists annotated the reports independently; one was designated the Reference Annotator (RA) and the other the Second Annotator (SA). We implemented a grammar-constrained Mistral-7B pipeline and evaluated its agreement with the RA alongside the SA and classical NLP baselines using Core Agreement and Certainty-Adjusted Agreement. A portability analysis held the task, labels, output grammar, report identifiers, reference annotations, and endpoints fixed while a MedGemma configuration used its model-declared instruction-turn format.

**Results:** For overall abnormality, Mistral-7B achieved 96% Core Agreement with the RA and the SA achieved 98%. Mistral-7B exceeded the tested low-resource base-lines in most categories and identified rare epileptiform abnormalities for which the baselines produced no or few true positives. Among reports with matching overall-abnormality decisions, Conditional Certainty Agreement was about 80% for Mistral–RA on each evaluated report set and 90% and 94% for SA–RA. The MedGemma configuration showed higher agreement than Mistral-7B in several non-epileptiform categories, but focal epileptiform performance involved a trade-off between missed and additional positive calls.

**Conclusions:** Under the studied report-level conditions, the grammar-constrained pipeline produced structured annotations with high binary agreement to the RA. After adapting the prompt presentation to its model-declared instruction-turn format, the same structured task and evaluation framework transferred to a newer local model. The results support its use as a research annotation method; they do not establish diagnostic ground truth, external validation, or readiness for clinical deployment.

**Significance:** This study demonstrates a locally executable method for converting de-identified EEG narratives into structured research annotations while retaining an ordinal representation of annotator certainty.

## I. Introduction

MODERN clinical neurophysiology generates vast volumes of diagnostic information, predominantly captured in unstructured, free-text narrative reports [1]. While essential for patient care, this narrative format creates a bottleneck for scalable analysis, quality assurance, and the training of machine learning models [2], [3]. Specifically, the variability in style and terminology, even among experts following American Clinical Neurophysiology Society (ACNS) guidelines, leads to interpretation ambiguity and hinders the systematic extraction of findings [4]. A persistent challenge remains the consistent capture of diagnostic uncertainty, often expressed through nuanced, implicit phrasing [5].

To address these issues, researchers have employed various Natural Language Processing (NLP) techniques. Early rule-based systems lacked flexibility [6], while transformer-based architectures like BERT improved classification by capturing contextual semantics [7], [8]. More recently, Large Language Models (LLMs) like Med-PalM2 have demonstrated high fluency in interpreting complex clinical narratives [9]. However, their application in high-stakes domains is constrained by concerns regarding factual accuracy and calibrated uncertainty [10], [11]. We therefore evaluate a framework that combines structured classification of EEG abnormalities with an ordinal representation of annotator certainty.

This study evaluates a grammar-constrained LLM pipeline designed to annotate free-text EEG reports. Our primary research question is: How closely do its structured labels agree with the study’s reference annotations for different abnormalities and certainty tiers? We expected higher agreement for the binary presence/absence decision than for an exact match on the four-level ordinal label.

We also explored the model’s ability to generate traceable justifications (explainability) and compared performance against various baseline models and LLMs. The description and evaluation of explanations, baseline model comparisons, and pipeline adaptations are detailed in the Supplementary Materials and further elaborated in [12].

## II. Methodology

### A. Data Source of Patients’ EEG reports and Cohort Selection

The study cohort was derived from a large corpus of EEG reports generated during routine clinical diagnostic workups at four hospitals in British Columbia, Canada. These reports, authored by multiple neurologists, were extracted from the electronic health record (EHR) system. The initial dataset represents a diverse collection of clinical language, reflecting variations in documentation practices, report structure, and descriptive terminology. To create a focused and manageable dataset for model development and evaluation, we selected two subsets of these reports for further processing and annotation.

The two subsets, referred to as the *Zoe Dataset* and the *Maria Dataset*, were authored by two different neurologists. The names “Zoe” and “Maria” are pseudonyms, used solely to distinguish reports produced by different neurologists. Each report was authored by one neurologist entirely. As shown in Table I, the two datasets vary in their length and vocabulary, showcasing the diversity of EEG reporting. The Supplementary Materials show sample reports from each neurologist. Reports from Zoe Dataset are more compact and consistent, while Maria’s reports have a broader vocabulary and are more variable in style. The Zoe Dataset serves as the primary dataset for development and evaluation. The Maria Dataset assesses transfer to reports written by a second neurologist within the same regional health authority; it is not an external-institution validation cohort.

**TABLE I:**
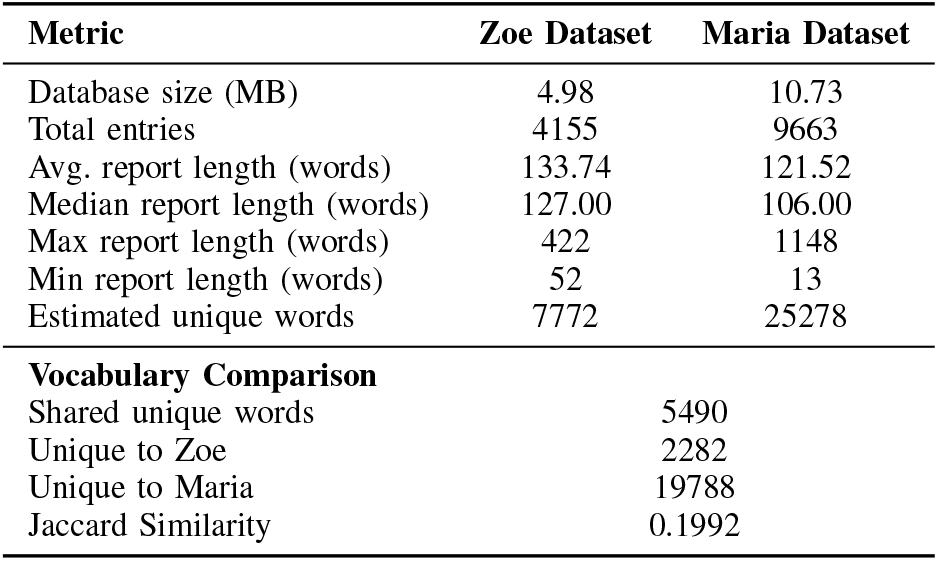
Characteristics of the source report collections for two neurologists, denoted by the pseudonyms Zoe and Maria. Values describe the available databases before study-sample selection; the annotated cohorts used for development and evaluation are reported in Table II. Jaccard similarity is the size of the shared vocabulary divided by the combined unique vocabulary.

### B. Annotation Schema

To implement the classification of clinical EEG reports, we develop a novel annotation schema. This schema enhances the classification framework proposed by Tveit et al. [13] by introducing new features: a four-point diagnostic confidence scale for each annotation. This section describes the schema’s hierarchical structure, category definitions, and rationale, which are designed to capture the complexity and uncertainty inherent in clinical EEG interpretation.

The annotation process follows a hierarchical structure to ensure the internal consistency of each labeled report. The initial step requires an overall assessment of the EEG report, classifying it as either normal or abnormal. This global classification constrains subsequent annotations. If the EEG in a report is classified as abnormal, annotators are required to select at least one of the four specific abnormality categories. Conversely, if the EEG in a report is classified as normal, annotators are instructed not to mark the presence of any abnormalities. This design ensures that every report labeled ’abnormal’ has an explicitly defined reason, while reports labeled ’normal’ are not associated with any conflicting abnormality markers.

Our schema permits the annotation of multiple, co-occurring abnormalities within a single report. This multi-label framework reflects clinical practice, which recognizes that patients may present with multiple distinct electrographic findings [14]. We instruct annotators to evaluate each report for the presence or absence of four specific categories of abnormal activity: focal epileptiform, generalized epileptiform, focal non-epileptiform, and generalized non-epileptiform. This approach is designed to produce a more complete and clinically representative summary of the EEG.

To quantify the certainty of each clinical judgment, we implement a four-point ordinal scale. For the overall assessment, annotators assign a score of 1 for a confident normal interpretation, 2 for a low-confidence normal interpretation, 3 for a low-confidence abnormal interpretation, or 4 for a confident abnormal interpretation. For each of the four abnormality categories, we apply a similar scale, where 1 indicates a confident absence, 2 indicates a low-confidence absence, 3 indicates a low-confidence presence, and 4 indicates a confident presence of the specific finding. This numerical system provides a structured method to capture the grader’s confidence alongside the primary clinical interpretation. The inclusion of this confidence metric serves two primary purposes. First, it enables the selective filtering of data for future model training. By creating training subsets that use only high-confidence annotations (scores of 1 or 4), we can reduce label noise and potentially improve the performance of a machine learning classifier. Second, the confidence scale provides a proxy for interpretative difficulty. We hypothesize that low-confidence annotations (scores of 2 or 3) correlate with EEG reports that exhibit high inter-rater variability among clinical experts. This creates a valuable dataset to investigate the sources of diagnostic uncertainty in neurophysiology.

### C. De-identification and Preprocessing Protocol

To prepare the clinical data for research while protecting patient privacy, we develop and apply a multistage deidentification protocol to each report. The primary objective of this protocol is to ensure the comprehensive removal of all Protected Health Information (PHI), ensuring compliance with institutional and ethical standards for data handling. The protocol involves three sequential steps.

First, we perform a systematic anonymization of structured identifiers. The original report and patient identifiers are replaced with unique hashed versions to maintain data linkage without exposing original information. Similarly, the names of the authoring neurologists are substituted with aliases to anonymize the source while preserving the ability to group reports by author. Standardized content found in report headers and footers, which often contains institutional or personal data, is also systematically removed.

Second, we employ a pattern-based redaction process to address PHI embedded within the narrative body of the reports. We use regular expression-based pattern matching to detect and remove common forms of PHI, including patient names, specific dates, telephone and fax numbers, email addresses, and hospital unit identifiers. This step targets predictable and consistently formatted sensitive information that might appear in the free-text portion of the reports.

Finally, to capture any residual PHI not identified by the preceding steps, we use a state-of-the-art NLP tool for a final anonymization scan. Each report is processed using Microsoft’s Presidio, an advanced analyzer and anonymizer designed to detect a wide range of sensitive entities within unstructured text [15]. This final step provides an additional layer of assurance, leveraging a sophisticated model to identify less common or irregularly formatted PHI. Only after a report successfully passes all three de-identification stages is it included in the final research corpus.

### D. Annotation Process and Rater Designation

The recovered study code used fixed source positions rather than random sampling. From 4,155 Zoe reports, positions 0– 499 and 1,000–1,999 yielded 1,495 complete annotated reports after five unusable records were excluded; the first 100 reports formed the prompt-development set and the remaining 1,395 the evaluation set. From 9,663 Maria reports, positions 0–499 yielded 499 complete annotated reports after one unusable record was excluded. All selected reports were independently annotated by two certified EEG technicians accredited by the Canadian Association of Electroneurophysiology Technologists (CAET). The Chief EEG Technician served as the *Reference Annotator (RA)* for agreement analyses. The second EEG Technician served as the *Second Annotator (SA)* for the human–human comparison. The RA labels are a study reference, not independently adjudicated diagnostic ground truth.

Both annotators independently applied the annotation schema described in Section II-B to selected reports in the Zoe and Maria datasets. The task required them to assess each report for the overall presence of abnormality as well as for four specific subtypes of abnormal activity. For each classification decision, annotators assigned a confidence level using a four-point ordinal scale, capturing both the diagnostic judgment and the degree of certainty associated with it. This process produced labels that reflect the clinical variability and interpretative confidence of expert readers. The label distributions for the Zoe dataset (*n* = 1,495) and the Maria dataset (*n* = 499), as annotated by the RA, are presented in Table II.

**TABLE II:** Reference Annotator (RA) label distribution in the selected reports, shown as count (percentage). Ordinal levels 1–2 are mapped to absence (No) and levels 3–4 to presence (Yes). Zoe combines 100 prompt-development and 1,395 evaluation reports; Maria contains 499 evaluation reports. Counts are report-level and do not establish patient independence.

| Category | Zoe ($n = 1,495$) | | Maria ($n = 499$) | |
| --- | --- | --- | --- | --- |
|  | No | Yes | No | Yes |
| Abnormality | 751 (50.2%) | 744 (49.8%) | 282 (56.5%) | 217 (43.5%) |
| Focal Epi | 1,437 (96.1%) | 58 (3.9%) | 467 (93.6%) | 32 (6.4%) |
| Gen Epi | 1,429 (95.6%) | 66 (4.4%) | 479 (96.0%) | 20 (4.0%) |
| Focal Non-Epi | 1,096 (73.3%) | 399 (26.7%) | 352 (70.5%) | 147 (29.5%) |
| Gen Non-Epi | 1,101 (73.6%) | 394 (26.4%) | 423 (84.8%) | 76 (15.2%) |

The four abnormality sub-categories are imbalanced in the selected reports. Focal and generalized epileptiform activity are present in approximately 4–6% of each report set. These small positive counts make category-specific estimates less precise. Because the reports were selected from fixed source positions, the observed proportions are not interpreted as population prevalence estimates.

### E. Proposed Method: A Grammar-Constrained LLM Pipeline

We selected Mistral-7B-Instruct-v0.2 as the core LLM for its reported performance and computational efficiency [16]. We used the Q5 K M quantized artifact [17] with temperature set to 0 on a single 12-GB NVIDIA GeForce RTX 3060 GPU. This configuration supported local execution without distributed infrastructure.

Our methodology employed a two-stage prompting pipeline, which is detailed in the Supplementary Materials and [12]. In the first stage, the prompt provided the LLM with the task definition and detailed annotation guidelines. The model was instructed to return classifications in a structured JSON format (Figure 1), ensuring predictions could be automatically parsed. To guarantee validity, we enforced a formal grammar constraint using the llama.cpp library [18], restricting the model’s generation to the predefined JSON schema.

**Fig. 1:**
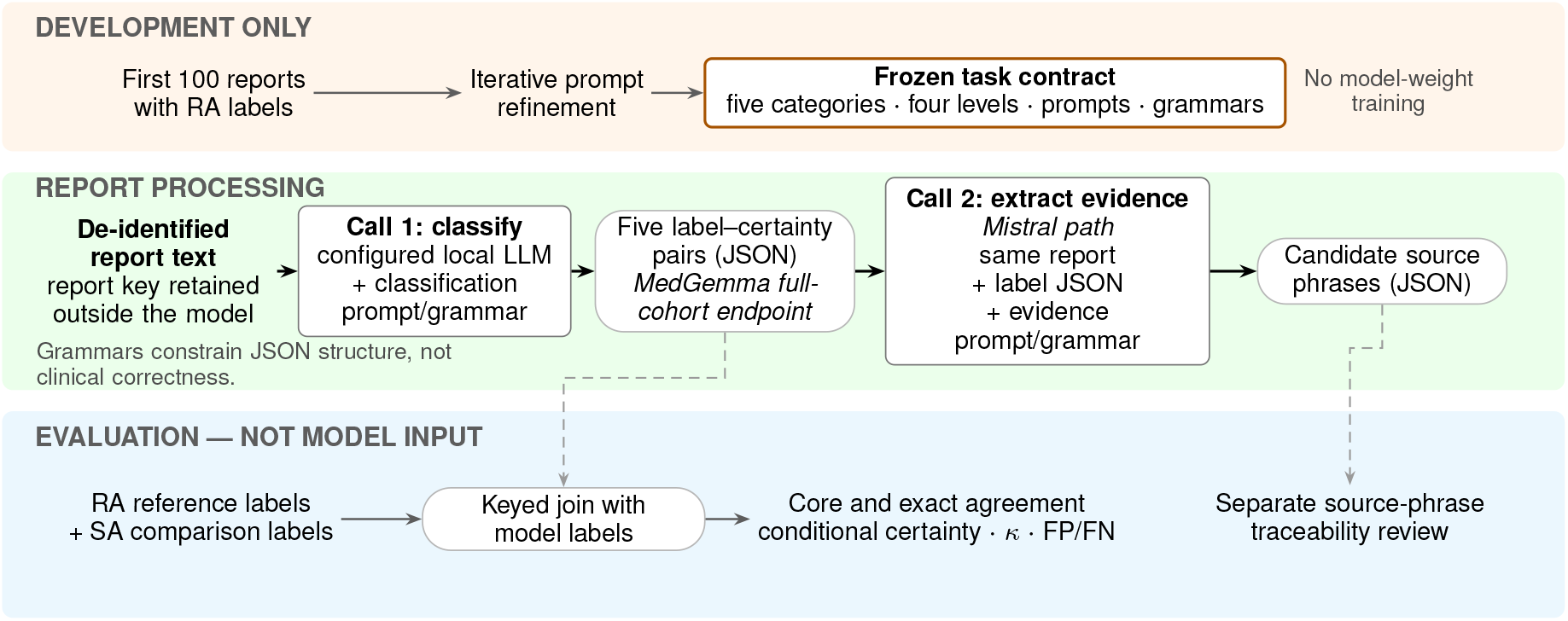
Development, report processing, and evaluation are separated. During development, the first 100 Zoe reports and Reference Annotator (RA) labels informed prompt refinement, not model-weight training. During report processing, each deidentified EEG report enters Call 1, which returns five label–certainty pairs under a constrained JSON grammar. The Mistral pathway then passes the report and labels to Call 2 to extract candidate source phrases; the full-cohort MedGemma comparison stops at the structured labels. RA labels and independent Second Annotator (SA) labels re-enter only after inference for report-keyed agreement analyses. Arrows show data flow rather than internal model reasoning, and the grammars constrain output structure, not clinical correctness.

In the second stage, a separate prompt asked the model to extract report phrases associated with each abnormal classification. These phrases provide candidates for source-traceability review; they are not treated as the model’s internal reasoning or as clinical adjudication.

For the MedGemma portability analysis, a model adapter placed the unchanged classification task and each de-identified EEG report inside the model-declared instruction-turn format expected by the pinned, locally cached MedGemma GGUF artifact [19]. Report keys, task definitions, five output categories, four-level labels, grammar, deterministic decoding, reference annotations, and evaluation endpoints remained fixed. All EEG report text remained on governed local storage, and inference used the local llama.cpp API; no report text was sent to a hosted service. This full-cohort analysis was classification-only and did not repeat Mistral’s second evidence-extraction call.

The prompt design was iteratively refined using the first 100 RA-annotated Zoe reports. Instructions were adjusted to improve clarity, incorporate definitions from established EEG reporting guidelines [20], and enforce consistency across related labels. The remaining 1,395 Zoe reports were reserved for evaluation; the recovered record does not contain a pre-specified prompt-development stopping rule.

#### Baseline Comparisons and Explanation Evaluation

To contextualize the performance of the proposed pipeline, we benchmarked it against two baseline architectures: Bag-of-Words with Logistic Regression (BoW+LR) and BERT with Logistic Regression (BERT+LR). The BoW+LR model served as a simple, transparent baseline reflective of traditional NLP techniques, while the BERT+LR model represented a context-aware transformer approach that remained computationally lightweight compared with the LLM [7], [21]. We used logistic regression for both baselines because of its efficiency and interpretability [22], [23]. We also retained feature-importance outputs for the baselines and candidate source phrases from the LLM’s second call. The Supplementary Materials describe these outputs and their traceability limits; they are not used as evidence of clinical correctness.

### F. Annotation Evaluation Framework

We compared automated annotations with the RA labels and used the independently produced SA labels as a human– human reference. Evaluations used 1,395 Zoe reports not used for prompt development and 499 Maria reports. Performance was assessed as Core Agreement (agreement on the binary presence/absence decision) and Certainty-Adjusted Agreement (exact agreement on the four-level ordinal label). We also report Cohen’s kappa to describe agreement beyond chance. These endpoints measure agreement with the study reference; they do not establish diagnostic correctness.

1. *Core Agreement:* Core Agreement assesses whether the model and RA agree on the presence or absence of each EEG finding, irrespective of the associated certainty tier. Labels 1 and 2 are collapsed into “Absence,” while labels 3 and 4 are collapsed into “Presence.” We report accuracy, precision, recall/sensitivity, specificity, and F1 score for each category [24], treating the RA label as the comparison reference.
2. *Certainty-Adjusted Agreement:* Certainty-Adjusted Agreement requires an exact match between a prediction and the RA label on the full four-point ordinal scale. It therefore combines agreement on the binary decision and the recorded certainty tier. We report the percentage of exact matches for each category. This endpoint is descriptive ordinal agreement, not a probability-calibration measure.
3. *Inter-rater Reliability:* To contextualize performance, we quantify agreement between the two human annotators and between each annotator and the evaluated systems. Cohen’s kappa (*κ*) describes observed agreement after accounting for agreement expected from the raters’ marginal label frequencies [25]. A value of 1 indicates perfect agreement and a value of 0 indicates agreement equal to that expected from those marginals. We calculated *κ* for each pairwise comparison under both Core and Certainty-Adjusted Agreement. Because kappa is affected by category prevalence and rater bias, we interpret it together with the confusion-matrix counts rather than as diagnostic accuracy.

## III. Results

### A. Core Agreement Performance

The Supplementary Materials detail Core Agreement on the primary Zoe dataset and the stylistically distinct Maria dataset. These analyses compare configured-system agreement with the RA across the two report-author surfaces.

On the Zoe dataset, the SA and Mistral-7B both showed high agreement with the RA for overall abnormality, with F1 scores of 0.98 and 0.96, respectively. The BoW+LR and BERT+LR baselines had F1 scores of 0.91 and 0.92, respectively, and produced fewer positive predictions.

The clearest differences occurred for focal and generalized epileptiform activity, the two categories with the fewest RA-positive reports. Seven evaluated baseline results had no true positives: BoW+LR for focal and generalized epileptiform activity on Zoe reports; BERT+LR for generalized epileptiform activity on Zoe reports; and both baselines for both epileptiform categories on Maria reports. On Zoe focal epileptiform activity, BERT+LR produced one true positive and 52 false negatives (F1 = 0.04), while BoW+LR produced no true positives. Mistral-7B and the SA each achieved an F1 score of 0.85 for focal epileptiform activity.

For focal non-epileptiform activity, Mistral-7B achieved an F1 score of 0.76, compared with 0.48 for BoW+LR and 0.57 for BERT+LR. For generalized non-epileptiform activity, the SA and Mistral-7B achieved F1 scores of 0.90 and 0.78, respectively, both higher than the tested baselines.

The Maria dataset tested the same configured systems on reports written by a second neurologist with distinct terminology within the same regional health authority. The SA remained highly concordant with the RA. Mistral-7B retained high Core Agreement in several categories, while the baseline results varied by category and report-author surface. These observations support within-authority transfer across the two selected reporting styles; they do not establish external or clinical generalizability.

### B. Certainty-Adjusted Agreement Performance

Core Agreement measures whether the binary presence/absence decision matches the RA, whereas Certainty-Adjusted Agreement requires an exact match on the four-point ordinal label. Because an exact four-level match necessarily includes a binary match, we additionally report Conditional Certainty Agreement: among reports with the same binary decision, the proportion with the same certainty tier. Equivalently, exact four-level agreement equals Core Agreement multiplied by Conditional Certainty Agreement. These quantities describe ordinal agreement; they are not probability-calibration measures.

The conditional decomposition makes the binary and certainty components separately interpretable (Figure 2). For overall abnormality, Mistral–RA certainty tiers matched in 1,077 of 1,341 binary matches (80.3%) on the Zoe reports and 363 of 457 (79.4%) on the Maria reports. The corresponding SA–RA values were 1,237 of 1,368 (90.4%) and 462 of 491 (94.1%).

**Fig. 2:**
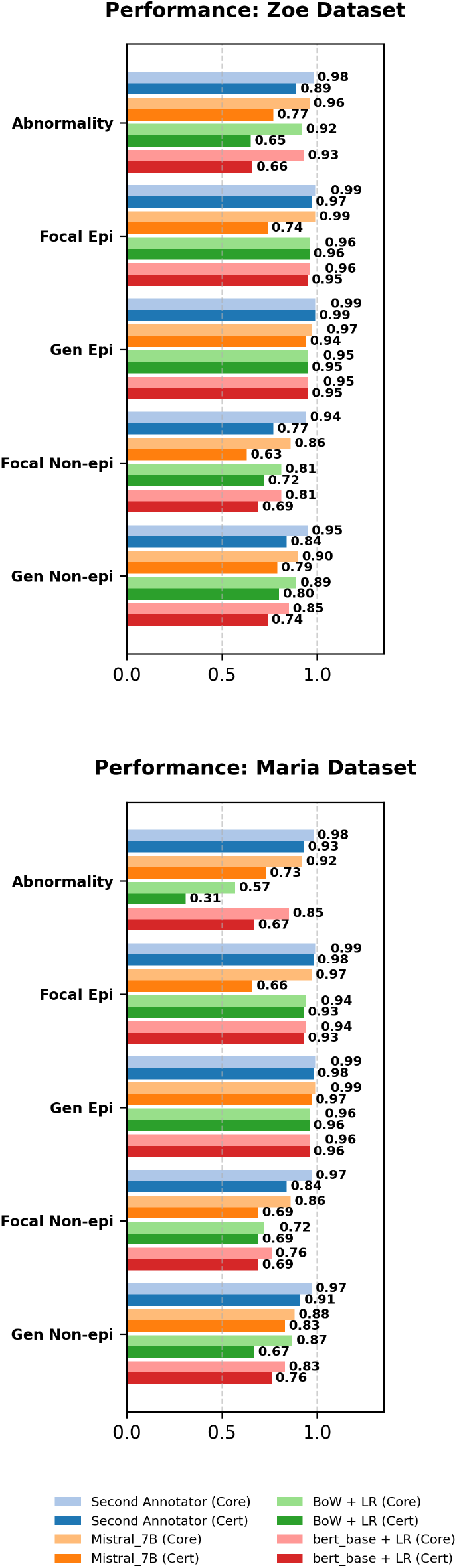
Agreement with the designated Reference Annotator (RA) on the 1,395 Zoe and 499 Maria evaluation reports. Core Agreement counts a match after ordinal levels 1–2 are mapped to absence and 3–4 to presence; Certainty-Adjusted Agreement requires the exact four-level label. SA denotes the Second Annotator; BoW+LR and BERT+LR are the two classical baselines. Values are report-level agreement with the RA, not diagnostic accuracy.

Category-specific results are reported in the Supplementary Materials, including separate denominators for jointly absent and jointly present decisions. This split is important for rare findings: a high aggregate conditional value can be dominated by the many reports that both raters marked absent. The decomposition describes certainty-tier concordance conditional on the binary decision; it does not establish probabilistic calibration, clinical correctness, a causal account of ambiguity, or external generalization.

### C. Inter-Rater Reliability Performance

To evaluate the consistency of the models’ classifications relative to human experts, we measure inter-rater reliability using Cohen’s Kappa (*κ*). This metric accounts for the probability of agreement occurring by chance, which is particularly important when dealing with the imbalanced class distributions found in clinical datasets [25]. We compute Kappa scores for each pair of raters (human-human, human-model, model-model) across all five diagnostic categories, first for core agreement and then for the more stringent certainty-adjusted agreement, for both the Zoe Dataset and the Maria Dataset (Figure 3).

**Fig. 3:**
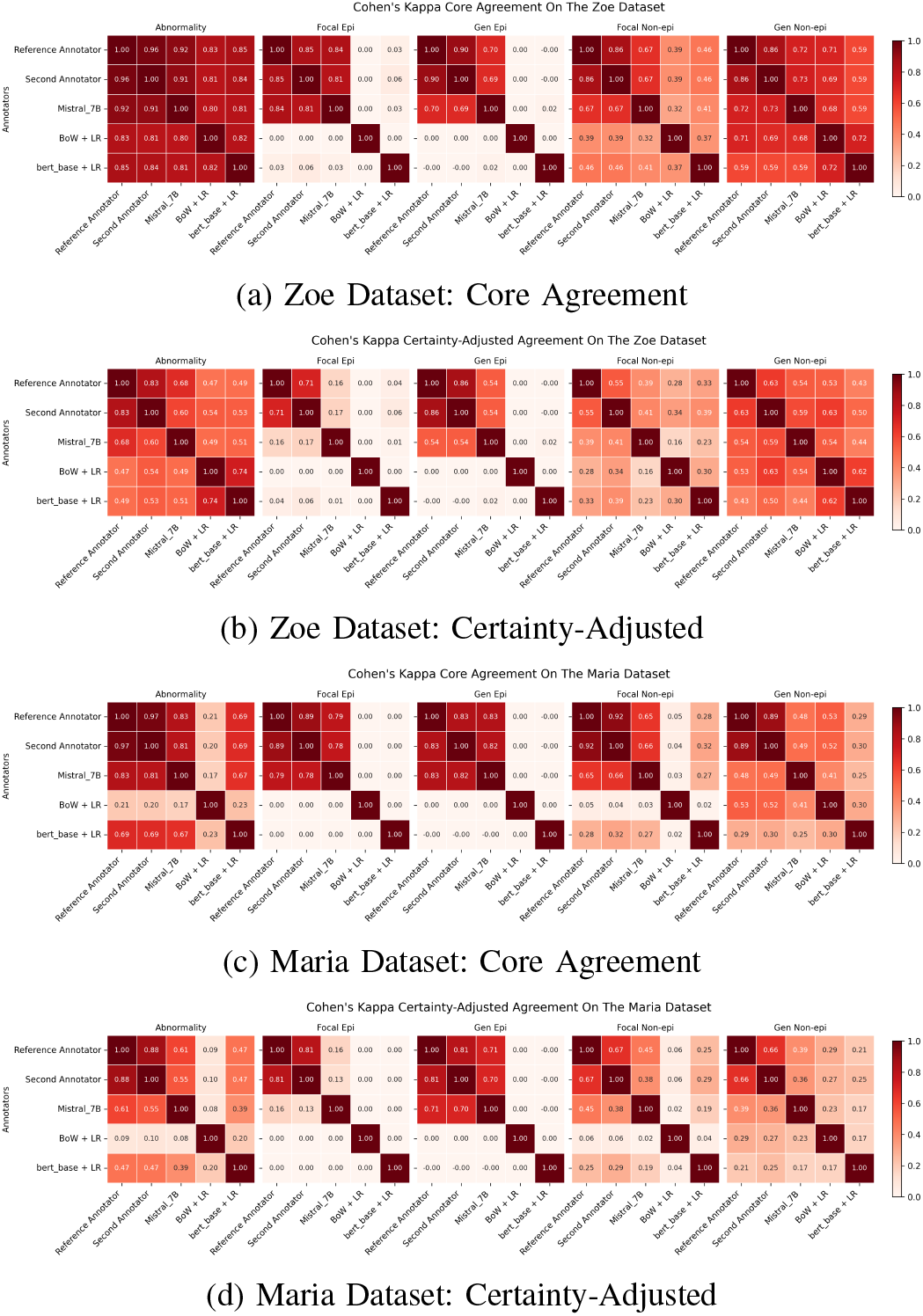
Pairwise chance-adjusted agreement among the Reference Annotator (RA), Second Annotator (SA), and evaluated systems. Heatmaps show Cohen’s *κ* on the 1,395 Zoe evaluation reports (a, b) and 499 Maria reports (c, d) for five categories. Panels (a) and (c) use Core Agreement after ordinal levels 1–2 are mapped to absence and 3–4 to presence; panels (b) and (d) require the exact four-level label. BoW+LR denotes Bag-of-Words plus logistic regression. Darker red indicates higher agreement. Kappa should be read with the RA-positive counts in Table II, because rare positive findings and differing marginal label frequencies affect its value; it is not diagnostic accuracy.

On the Zoe reports, RA–SA agreement was high for overall abnormality (*κ* = 0.96) and for focal and generalized epileptiform activity (*κ* = 0.85 and 0.90, respectively). Against the RA, Mistral-7B achieved *κ* = 0.92 for abnormality and *κ* = 0.84 for focal epileptiform activity. The tested baselines had near-zero kappa where they produced no or almost no positive predictions.

Under exact four-level agreement, kappa decreased across comparisons. RA–SA agreement remained highest for overall abnormality (*κ* = 0.83), while Mistral-7B kappa was 0.16 for focal epileptiform activity and 0.54 for generalized epileptiform activity. Maria results were category-specific. For BoW+LR abnormality, kappa was 0.21 under Core Agreement and 0.09 under exact four-level agreement. Kappa describes chance-adjusted agreement with the study annotations; it does not establish diagnostic accuracy, calibrated confidence, or clinical applicability.

## D. Contemporary-Model Portability Analysis

The MedGemma-27B Q2 K configuration produced valid structured classifications for the same 1,395 Zoe and 499 Maria evaluation reports, and every expected report key appeared exactly once. No report identifier or identical normalized report text appeared in both the 100-report development set and the 1,894-report evaluation set. Table III compares the Mistral-7B and MedGemma configured systems on the same reports under the same structured classification task and evaluation framework. Core Agreement increased most for non-epileptiform findings, but the result was not uniformly favorable. For Zoe focal epileptiform activity, MedGemma identified all 53 RA-positive reports, whereas Mistral missed two. Among 1,342 RA-negative reports, positive calls that disagreed with the RA increased from 16 (1.2%) to 36 (2.7%); precision consequently decreased from 76.1% to 59.6%, and Core Agreement from 98.7% to 97.4%. Here, “false positive” denotes disagreement with the RA, not an independently adjudicated clinical error.

**TABLE III:**
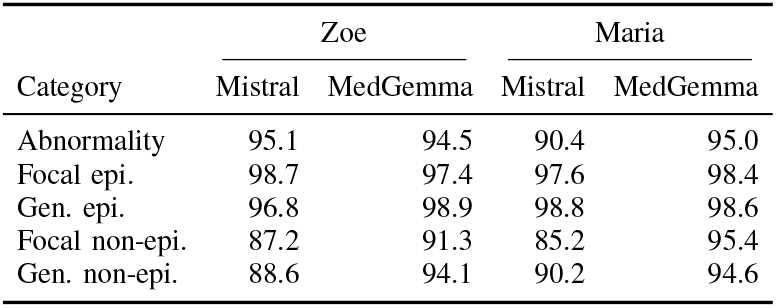
Same-report Core Agreement (%) with the Reference Annotator (RA) for the Mistral-7B and MedGemma configured systems on 1,395 Zoe and 499 Maria evaluation reports. Core Agreement is binary presence/absence agreement after mapping ordinal levels 1–2 to absence and 3–4 to presence. The task, labels, output grammar, reports, reference annotations, and evaluation endpoints were held fixed; MedGemma used its model-declared instruction-turn format. Values are agreement with the RA, not diagnostic accuracy.

| Category | Zoe |  | Maria |  |
| --- | --- | --- | --- | --- |
|  | Mistral | MedGemma | Mistral | MedGemma |
| Abnormality | 95.1 | 94.5 | 90.4 | 95.0 |
| Focal epi. | 98.7 | 97.4 | 97.6 | 98.4 |
| Gen. epi. | 96.8 | 98.9 | 98.8 | 98.6 |
| Focal non-epi. | 87.2 | 91.3 | 85.2 | 95.4 |
| Gen. non-epi. | 88.6 | 94.1 | 90.2 | 94.6 |

The supplementary material retains the complete paired statistics, the earlier lower-agreement evaluation using unmarked raw-completion input, and development-set same-model checks of input format. Those checks showed that MedGemma required its model-declared instruction-turn format for effective use in this pipeline, whereas the analogous 100-report Mistral check showed no consistent benefit. The output grammar remained active in both formats.

## IV. Discussion

### A. Summary and Interpretation of Findings

This study addressed the lack of structure in free-text EEG reports by combining a grammar-constrained LLM pipeline with a certainty-aware annotation schema. Mistral-7B showed high agreement with the RA on the binary finding decisions and generally exceeded the tested low-resource baselines. Conditional Certainty Agreement then isolates the nested question of whether the certainty tier also matched once the binary decision aligned. For overall abnormality, Mistral–RA certainty-tier concordance was stable at about 80% across the two report-author surfaces, while SA–RA concordance was 90–94%.

The prompt was developed iteratively on the first 100 Zoe reports using task definitions, examples, field ordering, consistency conditions, and endpoint guidance. The recovered record does not provide a prespecified stopping rule or a held-out ablation supporting the reported “up to 14%” causal improvement; that value is therefore omitted. The recovered component history still permits a bounded sensitivity analysis: in a newly specified one-factor replay on the same 100-report development set, removing only endpoint-confidence guidance reduced endpoint use by 4.4 percentage points and exact four-level agreement by 4.6 points, while binary agreement changed by 0.4 points. This shows that the instruction shaped ordinal output on that development set; it is not a reconstruction of the historical 14% result or evidence of held-out improvement. The Maria results describe transfer to a second neurologist’s reporting style within the same health authority, not external validation.

The second Mistral call extracted candidate source phrases after classification. These outputs provide reviewable evidence candidates but are not internal reasoning or clinical adjudication. Because the exact matcher used for the reported traceability percentage was not retained, no traceability percentage is reported, and explanation alignment is not used as evidence of clinical correctness or model self-awareness.

This analysis demonstrates portability of the structured task and evaluation framework across two configured local systems. The task contract and evaluation surface were held fixed; the model and its required input adapter changed. Because MedGemma and Mistral differ in model family, parameter count, and quantization, the comparison does not identify the effect of model weights alone. Category-specific gains and the Zoe focal-epileptiform sensitivity–precision trade-off are retained together. For research cohort construction, the additional positive calls that disagreed with the RA would increase the number of records requiring review; their clinical significance is not established. The same input-format intervention showed no consistent benefit in the smaller Mistral check, so an adapter effect should be validated for each configured artifact rather than assumed to transfer across models.

### B. Strengths of the Study

This study offers several methodological strengths. First, the structured annotation schema represents both abnormality and an ordinal certainty tier. Second, prompt guidance and constrained decoding produced machine-readable output on a quantized model using local hardware. Third, the independent RA and SA annotations expose human–human variability, while two neurologists’ report sets test the configured systems across differing writing styles within one health authority. Finally, the second model call retains candidate source phrases for traceability review, while keeping phrase extraction distinct from classification and clinical adjudication.

### C. Limitations of the Study

Reports came from one regional health authority and fixed source positions; the historical selection rationale and corpus representativeness are unknown. Maria represents a second neurologist’s reporting style, not an external institution. Analyses are report-level: a validated report-to-patient map was not available, so repeated-patient reports and patient overlap cannot be assessed and may make report-level intervals too narrow. Rare positive categories further limit precision. The RA is a study reference rather than diagnostic ground truth, and no adjudication of the full reference set is documented. Exact four-level agreement is not probabilistic calibration. Historical prompt stopping, baseline fold assignments, and fitted models were not fully retained. The Q5 K M result may depend on quantization and runtime. The MedGemma result likewise applies to one Q2 K configured system and cannot separate model-family, size, quantization, or input-format effects. No prospective workflow or clinical-benefit evaluation was performed; these limits preclude multicentre, clinical-utility, or deployment-readiness claims.

## A. Conclusion

This study shows that a locally executed, grammar-constrained Mistral-7B pipeline can convert free-text EEG reports into five structured ordinal annotations. On the selected report sets, its binary decisions showed high agreement with the RA and it identified rare epileptiform categories more successfully than the tested low-resource baselines. Exact agreement on the four-level labels remained lower.

The second report-author surface indicates that the configured pipeline can be applied across differing writing styles within the same regional health authority. The two-stage classify-then-extract design also retains candidate source phrases for later review. These findings support the framework as a reproducible research-annotation method. They do not establish diagnostic truth, calibrated confidence, external generalization, clinical efficiency, or readiness for clinical use.

## Data Availability

All data produced in the present study are available upon reasonable request to the authors

## Acknowledgments

We gratefully acknowledge the financial support of the National Research Council (NRC) of Canada under project DHGA-116-1. This work is also associated with Canadian Institutes of Health Research (CIHR) project grant application 487031. The joint Simon Fraser University/Fraser Health Research Ethics Board approved the study under H18-02728. The PI confirmed retrospective secondary use of identifiable information without individual consent under TCPS 2 (2022), Article 5.5A. Reports were processed using the three-stage deidentification procedure described above. This research was enabled in part by support provided by the Digital Research Alliance of Canada (alliancecan.ca).

