## Supplementary Materials for "Automated Interpretation of EEG Reports Using a Large Language Model with Structured Confidence Outputs"

### Supplementary Scope

This supplement retains the complete category-level performance surface, conditional certainty decomposition, support and error counts, baseline implementation details, independent Mistral reproduction, configured-system MedGemma comparisons, and the endpoint-guidance sensitivity analysis. It reports agreement with the designated Reference Annotator (RA), not diagnostic ground truth, and it keeps development analyses distinct from held-out evaluation. Identifying report text and report-level outputs are excluded.

### Supplementary Materials

#### A. Report-to-Annotation Workflow

The pipeline connects keyed report records, a local language model and the evaluation code; it neither trains the model nor makes a clinical decision. It (1) loads a report identifier and report text; (2) requests four-level labels for five finding categories (Prompt 1); (3) supplies those labels with the report to request supporting phrases (Prompt 2); and (4) retains keyed outputs and producing records for analysis. The identifier is not a patient name or a verified patient identifier, and it is not model input. Prompt 2 is decision-conditioned extraction, not a second independent classification or access to internal reasoning.

There is no conversation history from another report. Prompt 2 receives Prompt 1 labels explicitly, so the calls are sequential for a given report even when reports are batched. The decoding grammar restricts structure and permitted labels; it improves processing reliability, not correctness, verbatim quotation or clinical appropriateness. Source-present text still may not support the assigned category.

The structured task is evaluated as a configured local system: model artifact and quantization, input serialization, prompt, grammar, decoding, report population, reference annotations, and endpoints are named together. The MedGemma comparison holds the task, labels, output grammar, reports, reference, and endpoints fixed while using the model's declared instruction-turn format. Because model family, parameter count, and quantization also differ, the result tests operational portability of the framework rather than a base-model effect.

#### B. Supplementary Results: Table of Model Core Agreement Performance Comparison

This evidence layer compares the historical output lanes with the Reference Annotator (RA) while retaining the original report cohorts and endpoints. Table S1 gives the complete performance surface; the independent Mistral reproduction and configured MedGemma comparisons appear separately later. The table reports reference agreement, not explanation quality or diagnostic truth, and Maria is a second report-author surface within the same regional health authority rather than external institutional validation.

#### C. Conditional Certainty Agreement

The four-level label contains two nested questions: whether the finding is present or absent, and whether that decision is confident or low-confidence. Core Agreement answers the first question. Exact four-level agreement answers both simultaneously. We therefore add a descriptive decomposition, *Conditional Certainty Agreement*, defined as $E/B$, where $B$ is the number of binary matches and $E$ is the number of exact four-level matches. It asks: among reports for which a comparator and the RA made the same binary decision, how often did they also assign the same certainty tier? Equivalently, exact four-level agreement equals Core Agreement multiplied by Conditional Certainty Agreement. This identity explains the nesting; it is not a new model endpoint, probability calibration, or independently adjudicated correctness.

Table S2 reports exact paired counts calculated from the retained labels. The jointly absent and jointly present columns keep rare-category denominators visible. Without this split, a comparator that rarely assigns a positive label can appear to have high certainty agreement because nearly all of its binary matches are jointly absent. The low-resource baselines are therefore not ranked by this derived measure here; their binary detection failures remain visible in Tables S1 and S4.

For overall abnormality, Mistral–RA Conditional Certainty Agreement is stable across the two report-author surfaces

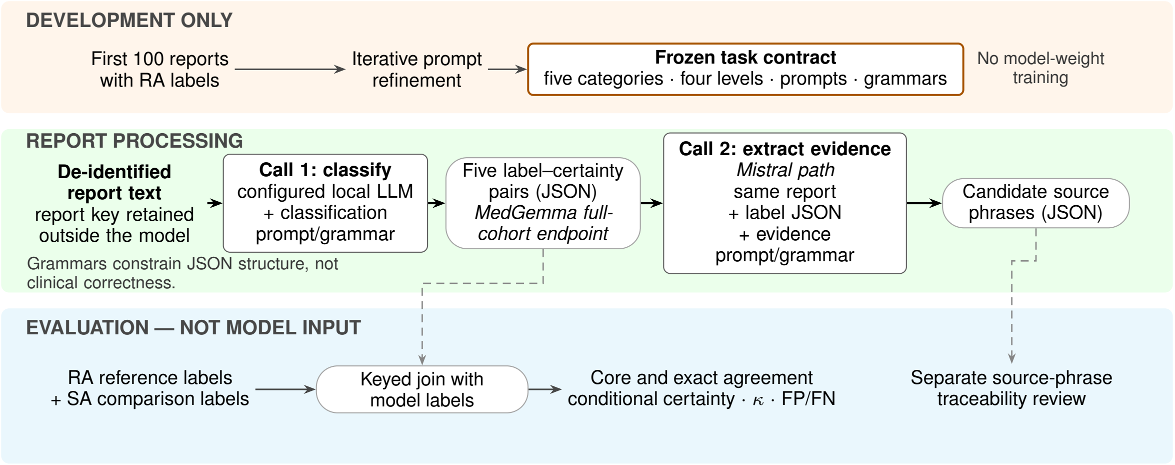


Fig. S1: **Development, report processing, and evaluation are separated.** During development, the first 100 Zoe reports and Reference Annotator (RA) labels informed prompt refinement, not model-weight training. During report processing, each de-identified EEG report enters Call 1, which returns five label–certainty pairs under a constrained JSON grammar. The Mistral pathway then passes the report and labels to Call 2 to extract candidate source phrases; the full-cohort MedGemma comparison stops at the structured labels. RA labels and independent Second Annotator (SA) labels re-enter only after inference for report-keyed agreement analyses. Arrows show data flow rather than internal model reasoning, and the grammars constrain output structure, not clinical correctness.

(80.3% and 79.4%), while SA–RA provides human–human context (90.4% and 94.1%). The category and stratum results are less uniform. Mistral–RA focal-epileptiform certainty tiers match in only 26/51 (51.0%) and 13/27 (48.1%) jointly present decisions on Zoe and Maria, respectively. Human–human certainty disagreement is also category-dependent: for Zoe focal non-epileptiform activity, SA and RA match tiers in 166/358 (46.4%) jointly present decisions. These findings make the certainty layer interpretable without treating the RA label as diagnostic truth.

#### D. Development and Evaluation Support Counts

This cohort-accounting layer fixes the denominators behind every later comparison. Table S3 reports report counts after levels 1–2 are mapped to core absence and 3–4 to core presence; RA and SA denote the Reference and Second Annotators, and the SA counts use the same complete reports as the RA analysis. All 1,994 study inputs matched both original annotator exports by report hash and text, with no missing keys or discrepancies. These counts support report-level interpretation but do not establish patient independence, source-corpus prevalence or clinical adjudication.

There are few reference-positive development reports. Generalized epileptiform activity has three RA-positive development reports, which cannot populate all five test folds; recall cannot be estimated in every fold. Focal epileptiform activity has exactly five RA-positive development reports. These small counts limit later confidence analysis and must remain visible rather than being hidden by an average or a category-specific fitted model.

#### E. Submitted Binary Error Counts

This submitted-error layer makes the counts behind Core Agreement visible. Table S4 uses the Reference Annotator as the comparison reference; $P$ and $N$ are RA-positive and RA-negative support after levels 1–2 are mapped to absence and 3–4 to presence. It supports category-specific review of misses and additional positive calls, while Cohen's $\kappa$ summarizes agreement beyond that expected from the two label frequencies. It remains an agreement audit, not an estimate of diagnostic truth or clinical harm.

The ten Zoe BoW+LR and BERT+LR matrices were recoverable only as hash-verified submitted aggregates. They support the displayed counts and point metrics but not new paired report-level intervals or case review. Matching keyed sources are retained under controlled access for the other 30 matrices.

Seven baseline/category surfaces have zero true positives: Zoe BoW+LR focal and generalized epileptiform activity, Zoe BERT+LR generalized epileptiform activity, and both Maria baselines for both epileptiform categories. Zoe BERT+LR focal epileptiform activity has one true positive and 52 false negatives. These unfavorable results are retained explicitly. Clinical severity, patient-level clustering, and the consequences of individual false negatives require qualified clinical case review and are not inferred from aggregate counts.

#### F. Baseline Models for Comparison

The submitted comparison used Bag-of-Words with Logistic Regression (**BoW+LR**) and BERT with Logistic Regression (**BERT+LR**) [1], [2]. Separate classifiers were fit for each binary outcome after mapping ordinal levels 1–2 to absence and 3–4 to presence.

The BoW representation used raw word/phrase counts: at most 10,000 features, with common English words removed (stop-word removal), sequences of one to five words (1–5-grams), and alphabetic tokens of at least three characters. Logistic regression learns a weight for each feature. It captures order within these short sequences, not across the whole report.

TABLE S1: **Model Core Agreement Performance Comparison across diagnostic categories for the Zoe and Maria EEG datasets.** The table reports agreement with the RA for the SA, Bag-of-Words plus Logistic Regression (BoW+LR), BERT plus Logistic Regression (BERT+LR), and Mistral-7B. Metrics include accuracy, precision, recall/sensitivity, F1, and specificity. Values are grouped by report author and five diagnostic categories. They describe the tested models on these selected report-level surfaces and do not establish diagnostic ground truth or external generalizability.

| Category | Metric | Second Annotator | BoW + LR | BERT + LR | Mistral-7B |
|---|---|---|---|---|---|
| | | **Zoe Dataset** | | | |
| Abnormality | Core Accuracy | 0.98 | 0.92 | 0.93 | 0.96 |
| | Core Precision | 0.99 | 0.98 | 0.95 | 0.97 |
| | Core Recall(Sensitivity) | 0.98 | 0.85 | 0.90 | 0.95 |
| | Core F1 Score | 0.98 | 0.91 | 0.92 | 0.96 |
| | Specificity | 0.99 | 0.98 | 0.95 | 0.97 |
| Focal Epi | Core Accuracy | 0.99 | 0.96 | 0.96 | 0.99 |
| | Core Precision | 0.79 | 0.00 | 0.50 | 0.76 |
| | Core Recall (Sensitivity) | 0.92 | 0.00 | 0.02 | 0.96 |
| | Core F1 Score | 0.85 | 0.00 | 0.04 | 0.85 |
| | Specificity | 0.99 | 1.00 | 1.00 | 0.99 |
| Gen Epi | Core Accuracy | 0.99 | 0.95 | 0.95 | 0.97 |
| | Core Precision | 0.95 | 0.00 | 0.00 | 0.60 |
| | Core Recall(Sensitivity) | 0.86 | 0.00 | 0.00 | 0.89 |
| | Core F1 Score | 0.90 | 0.00 | 0.00 | 0.71 |
| | Specificity | 1.00 | 1.00 | 1.00 | 0.97 |
| Focal Non-epi | Core Accuracy | 0.94 | 0.81 | 0.81 | 0.86 |
| | Core Precision | 0.86 | 0.89 | 0.74 | 0.72 |
| | Core Recall (Sensitivity) | 0.95 | 0.33 | 0.47 | 0.81 |
| | Core F1 Score | 0.90 | 0.48 | 0.57 | 0.76 |
| | Specificity | 0.94 | 0.98 | 0.94 | 0.88 |
| Gen Non-epi | Core Accuracy | 0.95 | 0.89 | 0.85 | 0.90 |
| | Core Precision | 0.92 | 0.86 | 0.76 | 0.90 |
| | Core Recall (Sensitivity) | 0.88 | 0.71 | 0.63 | 0.69 |
| | Core F1 Score | 0.90 | 0.78 | 0.69 | 0.78 |
| | Specificity | 0.97 | 0.96 | 0.93 | 0.97 |
| | | **Maria Dataset** | | | |
| Abnormality | Core Accuracy | 0.98 | 0.57 | 0.85 | 0.92 |
| | Core Precision | 0.99 | 0.50 | 0.80 | 0.95 |
| | Core Recall (Sensitivity) | 0.97 | 1.00 | 0.87 | 0.85 |
| | Core F1 Score | 0.98 | 0.67 | 0.83 | 0.90 |
| | Core Specificity | 0.99 | 0.23 | 0.83 | 0.97 |
| Focal Epi | Core Accuracy | 0.99 | 0.94 | 0.94 | 0.97 |
| | Core Precision | 0.96 | 0.00 | 0.00 | 0.77 |
| | Core Recall (Sensitivity) | 0.84 | 0.00 | 0.00 | 0.84 |
| | Core F1 Score | 0.90 | 0.00 | 0.00 | 0.81 |
| | Core Specificity | 1.00 | 1.00 | 1.00 | 0.98 |
| Gen Epi | Core Accuracy | 0.99 | 0.96 | 0.96 | 0.99 |
| | Core Precision | 0.78 | 0.00 | 0.00 | 0.78 |
| | Core Recall (Sensitivity) | 0.90 | 0.00 | 0.00 | 0.90 |
| | Core F1 Score | 0.84 | 0.00 | 0.00 | 0.84 |
| | Core Specificity | 0.99 | 1.00 | 1.00 | 0.99 |
| Focal Non-epi | Core Accuracy | 0.97 | 0.72 | 0.76 | 0.86 |
| | Core Precision | 0.92 | 1.00 | 0.73 | 0.80 |
| | Core Recall (Sensitivity) | 0.97 | 0.03 | 0.27 | 0.69 |
| | Core F1 Score | 0.94 | 0.07 | 0.40 | 0.74 |
| | Core Specificity | 0.97 | 1.00 | 0.96 | 0.93 |
| Gen Non-epi | Core Accuracy | 0.97 | 0.87 | 0.83 | 0.88 |
| | Core Precision | 0.94 | 0.55 | 0.43 | 0.68 |
| | Core Recall (Sensitivity) | 0.87 | 0.68 | 0.36 | 0.45 |
| | Core F1 Score | 0.90 | 0.61 | 0.39 | 0.54 |
| | Core Specificity | 0.99 | 0.90 | 0.91 | 0.96 |

The BERT baseline used `bert-base-uncased` to turn each report into a numerical feature vector (the final-layer [CLS] embedding). BERT's weights were kept fixed, not fine-tuned; text beyond 512 model tokens was cut off. Logistic regression used these features with `max_iter=1000`. The transferred files do not retain the exact training/test fold assignments, fitted classifiers, or random seed needed to replay training exactly.

**BERT input fairness: did end truncation change the comparison?**

The exact historical tokenizer and 512-token right-truncation rule exposed 1/1,395 Zoe reports (0.072%) and 3/499 Maria reports (0.601%). All four contained a recognized downstream heading. Two Maria reports had at least one Impression, Interpretation, Conclusion, or Clinical Correlation heading beyond the retained boundary; in one, the first recognized Interpretation heading itself began beyond it. Removing all four reports from *every* saved model surface changed no category-level Core Agreement ranking. For BERT+LR, the largest change was 0.106 percentage points in Core Agreement and 0.310 points in F1. This confirms that late-section truncation occurred, while bounding its observed influence on the saved

TABLE S2: Conditional Certainty Agreement with the RA from exact paired labels. $N$ is the report count, $B$ the number of matching binary decisions, and $E$ the number that also match exactly on the four-level label. Absent and present columns condition further on jointly absent and jointly present decisions. Percentages describe agreement with the designated RA, not clinical correctness or probability calibration.

| Report set | Comparator | Category | $B/N$ | $E/B$ | Jointly absent $E/B$ | Jointly present $E/B$ |
|---|---|---|---|---|---|---|
| Zoe evaluation | Mistral–RA | Abnormality | 1,341/1,395 | 1,077/1,341 (80.3%) | 578/675 (85.6%) | 499/666 (74.9%) |
| | Mistral–RA | Focal epi. | 1,377/1,395 | 1,028/1,377 (74.7%) | 1,002/1,326 (75.6%) | 26/51 (51.0%) |
| | Mistral–RA | Gen. epi. | 1,350/1,395 | 1,312/1,350 (97.2%) | 1,262/1,294 (97.5%) | 50/56 (89.3%) |
| | Mistral–RA | Focal non-epi. | 1,204/1,395 | 882/1,204 (73.3%) | 718/899 (79.9%) | 164/305 (53.8%) |
| | Mistral–RA | Gen. non-epi. | 1,254/1,395 | 1,108/1,254 (88.4%) | 930/999 (93.1%) | 178/255 (69.8%) |
| | SA–RA | Abnormality | 1,368/1,395 | 1,237/1,368 (90.4%) | 593/684 (86.7%) | 644/684 (94.2%) |
| | SA–RA | Focal epi. | 1,378/1,395 | 1,360/1,378 (98.7%) | 1,320/1,329 (99.3%) | 40/49 (81.6%) |
| | SA–RA | Gen. epi. | 1,383/1,395 | 1,379/1,383 (99.7%) | 1,328/1,329 (99.9%) | 51/54 (94.4%) |
| | SA–RA | Focal non-epi. | 1,317/1,395 | 1,080/1,317 (82.0%) | 914/959 (95.3%) | 166/358 (46.4%) |
| | SA–RA | Gen. non-epi. | 1,320/1,395 | 1,167/1,320 (88.4%) | 955/998 (95.7%) | 212/322 (65.8%) |
| Maria evaluation | Mistral–RA | Abnormality | 457/499 | 363/457 (79.4%) | 215/273 (78.8%) | 148/184 (80.4%) |
| | Mistral–RA | Focal epi. | 486/499 | 330/486 (67.9%) | 317/459 (69.1%) | 13/27 (48.1%) |
| | Mistral–RA | Gen. epi. | 492/499 | 485/492 (98.6%) | 467/474 (98.5%) | 18/18 (100.0%) |
| | Mistral–RA | Focal non-epi. | 429/499 | 343/429 (80.0%) | 267/327 (81.7%) | 76/102 (74.5%) |
| | Mistral–RA | Gen. non-epi. | 441/499 | 416/441 (94.3%) | 393/407 (96.6%) | 23/34 (67.6%) |
| | SA–RA | Abnormality | 491/499 | 462/491 (94.1%) | 261/280 (93.2%) | 201/211 (95.3%) |
| | SA–RA | Focal epi. | 493/499 | 488/493 (99.0%) | 465/466 (99.8%) | 23/27 (85.2%) |
| | SA–RA | Gen. epi. | 492/499 | 491/492 (99.8%) | 473/474 (99.8%) | 18/18 (100.0%) |
| | SA–RA | Focal non-epi. | 482/499 | 419/482 (86.9%) | 330/340 (97.1%) | 89/142 (62.7%) |
| | SA–RA | Gen. non-epi. | 485/499 | 454/485 (93.6%) | 414/419 (98.8%) | 40/66 (60.6%) |

TABLE S3: Aggregate support by cohort, annotator, category, and ordinal level. Counts are report-level and describe the evaluated reference surface; they are not patient counts.

| Cohort | Rater | Category | L1 | L2 | L3 | L4 | Absent | Present | N |
|---|---|---|---|---|---|---|---|---|---|
| Zoe development | RA | Abnormality | 50 | 7 | 20 | 23 | 57 | 43 | 100 |
| | RA | Focal epi. | 94 | 1 | 1 | 4 | 95 | 5 | 100 |
| | RA | Gen. epi. | 96 | 1 | 0 | 3 | 97 | 3 | 100 |
| | RA | Focal non-epi. | 68 | 11 | 10 | 11 | 79 | 21 | 100 |
| | RA | Gen. non-epi. | 72 | 2 | 10 | 16 | 74 | 26 | 100 |
| | SA | Abnormality | 51 | 4 | 10 | 35 | 55 | 45 | 100 |
| | SA | Focal epi. | 93 | 0 | 2 | 5 | 93 | 7 | 100 |
| | SA | Gen. epi. | 98 | 0 | 1 | 1 | 98 | 2 | 100 |
| | SA | Focal non-epi. | 71 | 2 | 6 | 21 | 73 | 27 | 100 |
| | SA | Gen. non-epi. | 75 | 1 | 0 | 24 | 76 | 24 | 100 |
| Zoe evaluation | RA | Abnormality | 573 | 121 | 339 | 362 | 694 | 701 | 1,395 |
| | RA | Focal epi. | 1,335 | 7 | 10 | 43 | 1,342 | 53 | 1,395 |
| | RA | Gen. epi. | 1,330 | 2 | 7 | 56 | 1,332 | 63 | 1,395 |
| | RA | Focal non-epi. | 955 | 62 | 229 | 149 | 1,017 | 378 | 1,395 |
| | RA | Gen. non-epi. | 975 | 52 | 158 | 210 | 1,027 | 368 | 1,395 |
| | SA | Abnormality | 678 | 23 | 316 | 378 | 701 | 694 | 1,395 |
| | SA | Focal epi. | 1,328 | 5 | 7 | 55 | 1,333 | 62 | 1,395 |
| | SA | Gen. epi. | 1,337 | 1 | 3 | 54 | 1,338 | 57 | 1,395 |
| | SA | Focal non-epi. | 957 | 22 | 28 | 388 | 979 | 416 | 1,395 |
| | SA | Gen. non-epi. | 1,034 | 10 | 33 | 318 | 1,044 | 351 | 1,395 |
| Maria evaluation | RA | Abnormality | 262 | 20 | 80 | 137 | 282 | 217 | 499 |
| | RA | Focal epi. | 466 | 1 | 3 | 29 | 467 | 32 | 499 |
| | RA | Gen. epi. | 478 | 1 | 0 | 20 | 479 | 20 | 499 |
| | RA | Focal non-epi. | 339 | 13 | 60 | 87 | 352 | 147 | 499 |
| | RA | Gen. non-epi. | 417 | 6 | 32 | 44 | 423 | 76 | 499 |
| | SA | Abnormality | 282 | 4 | 73 | 140 | 286 | 213 | 499 |
| | SA | Focal epi. | 470 | 1 | 4 | 24 | 471 | 28 | 499 |
| | SA | Gen. epi. | 476 | 0 | 0 | 23 | 476 | 23 | 499 |
| | SA | Focal non-epi. | 342 | 3 | 10 | 144 | 345 | 154 | 499 |
| | SA | Gen. non-epi. | 429 | 0 | 0 | 70 | 429 | 70 | 499 |

comparison. It is not a counterfactual long-context BERT run or proof of model equivalence.

For both baseline models, the continuous probability output ($\hat{p}$) from the logistic regression classifier was mapped to our four-point ordinal confidence scale using predefined thresholds. These thresholds, tuned on a development set of 100 RA-annotated reports from the Zoe dataset, were designed to capture uncertainty around the 0.5 decision boundary:

- $\hat{p} < 0.4$: Confident No (1)
- $0.4 \leq \hat{p} < 0.5$: Low Confidence No (2)
- $0.5 \leq \hat{p} < 0.6$: Low Confidence Yes (3)
- $\hat{p} \geq 0.6$: Confident Yes (4)

The historical implementation describes five-fold cross-validation on the 100-report Zoe development set: divide reports into five parts, train on four and assess the remaining part, rotating the held-out part. Evaluation on the larger report sets followed. Because the exact historical splits and fitted models are unavailable, we cannot reconstruct variation across folds or verify how the original split prevented training information from reaching test cases (data leakage). The four-level bins above are thresholded logistic-regression probabilities; they do not show that LLM confidence matches observed correctness (probability calibration), or establish clinical certainty.

### G. Reproducibility and Configured-System Evaluation

This protocol keeps submitted outputs, independent reproduction, configured-system comparisons, and development sensitivity analyses as separate evidence layers. Each layer retains its cohort, model, prompt, predictions, analysis, and checks; numerical results come from saved predictions and

TABLE S4: Submitted binary error counts against the RA. TP, true positive; FN, false negative; FP, false positive; TN, true negative. Error labels denote disagreement with the RA, not independently adjudicated clinical error.

| Cohort | Comparator | Category | P | N | TP | FN | FP | TN | $\kappa$ |
|---|---|---|---|---|---|---|---|---|---|
| Zoe | SA | Abnormality | 701 | 694 | 684 | 17 | 10 | 684 | 0.961 |
| Zoe | SA | Focal epi. | 53 | 1,342 | 49 | 4 | 13 | 1,329 | 0.846 |
| Zoe | SA | Gen. epi. | 63 | 1,332 | 54 | 9 | 3 | 1,329 | 0.896 |
| Zoe | SA | Focal non-epi. | 378 | 1,017 | 358 | 20 | 58 | 959 | 0.863 |
| Zoe | SA | Gen. non-epi. | 368 | 1,027 | 322 | 46 | 29 | 998 | 0.860 |
| Zoe | BoW+LR | Abnormality | 701 | 694 | 595 | 106 | 12 | 682 | 0.831 |
| Zoe | BoW+LR | Focal epi. | 53 | 1,342 | 0 | 53 | 0 | 1,342 | 0.000 |
| Zoe | BoW+LR | Gen. epi. | 63 | 1,332 | 0 | 63 | 0 | 1,332 | 0.000 |
| Zoe | BoW+LR | Focal non-epi. | 378 | 1,017 | 124 | 254 | 16 | 1,001 | 0.389 |
| Zoe | BoW+LR | Gen. non-epi. | 368 | 1,027 | 263 | 105 | 44 | 983 | 0.710 |
| Zoe | BERT+LR | Abnormality | 701 | 694 | 633 | 68 | 35 | 659 | 0.852 |
| Zoe | BERT+LR | Focal epi. | 53 | 1,342 | 1 | 52 | 1 | 1,341 | 0.034 |
| Zoe | BERT+LR | Gen. epi. | 63 | 1,332 | 0 | 63 | 2 | 1,330 | -0.003 |
| Zoe | BERT+LR | Focal non-epi. | 378 | 1,017 | 177 | 201 | 61 | 956 | 0.462 |
| Zoe | BERT+LR | Gen. non-epi. | 368 | 1,027 | 231 | 137 | 71 | 956 | 0.593 |
| Zoe | Mistral-7B | Abnormality | 701 | 694 | 666 | 35 | 19 | 675 | 0.923 |
| Zoe | Mistral-7B | Focal epi. | 53 | 1,342 | 51 | 2 | 16 | 1,326 | 0.843 |
| Zoe | Mistral-7B | Gen. epi. | 63 | 1,332 | 56 | 7 | 38 | 1,294 | 0.697 |
| Zoe | Mistral-7B | Focal non-epi. | 378 | 1,017 | 305 | 73 | 118 | 899 | 0.666 |
| Zoe | Mistral-7B | Gen. non-epi. | 368 | 1,027 | 255 | 113 | 28 | 999 | 0.719 |
| Maria | SA | Abnormality | 217 | 282 | 211 | 6 | 2 | 280 | 0.967 |
| Maria | SA | Focal epi. | 32 | 467 | 27 | 5 | 1 | 466 | 0.894 |
| Maria | SA | Gen. epi. | 20 | 479 | 18 | 2 | 5 | 474 | 0.830 |
| Maria | SA | Focal non-epi. | 147 | 352 | 142 | 5 | 12 | 340 | 0.919 |
| Maria | SA | Gen. non-epi. | 76 | 423 | 66 | 10 | 4 | 419 | 0.888 |
| Maria | BoW+LR | Abnormality | 217 | 282 | 217 | 0 | 217 | 65 | 0.207 |
| Maria | BoW+LR | Focal epi. | 32 | 467 | 0 | 32 | 0 | 467 | 0.000 |
| Maria | BoW+LR | Gen. epi. | 20 | 479 | 0 | 20 | 0 | 479 | 0.000 |
| Maria | BoW+LR | Focal non-epi. | 147 | 352 | 5 | 142 | 0 | 352 | 0.047 |
| Maria | BoW+LR | Gen. non-epi. | 76 | 423 | 52 | 24 | 42 | 381 | 0.533 |
| Maria | BERT+LR | Abnormality | 217 | 282 | 189 | 28 | 48 | 234 | 0.693 |
| Maria | BERT+LR | Focal epi. | 32 | 467 | 0 | 32 | 0 | 467 | 0.000 |
| Maria | BERT+LR | Gen. epi. | 20 | 479 | 0 | 20 | 1 | 478 | -0.004 |
| Maria | BERT+LR | Focal non-epi. | 147 | 352 | 40 | 107 | 15 | 337 | 0.281 |
| Maria | BERT+LR | Gen. non-epi. | 76 | 423 | 27 | 49 | 36 | 387 | 0.291 |
| Maria | Mistral-7B | Abnormality | 217 | 282 | 184 | 33 | 9 | 273 | 0.827 |
| Maria | Mistral-7B | Focal epi. | 32 | 467 | 27 | 5 | 8 | 459 | 0.792 |
| Maria | Mistral-7B | Gen. epi. | 20 | 479 | 18 | 2 | 5 | 474 | 0.830 |
| Maria | Mistral-7B | Focal non-epi. | 147 | 352 | 102 | 45 | 25 | 327 | 0.649 |
| Maria | Mistral-7B | Gen. non-epi. | 76 | 423 | 34 | 42 | 16 | 407 | 0.476 |

deterministic analysis rather than generated explanations. Development results remain non-independent, and missing historical producing files limit retrospective reconstruction.

**A fair comparison starts by naming the whole evidence surface**

A result is identified by the model artifact and quantization, input serialization, prompt, grammar, decoding, report population, reference annotator, endpoint and selection role. A controlled ablation changes one of these factors. A model-declared-serialization comparison holds the task, reports, reference and endpoints fixed while each local instruction-tuned model receives its receipted turn format. A configured-system comparison may change several factors and is useful operationally, but cannot isolate a base-model effect.

**Population crosswalk.** The reproduction processed 2,500 reports: 2,000 Zoe plus 500 Maria. The study-analysis population contains 1,994 usable reports: 100 Zoe development, 1,395 Zoe evaluation and 499 Maria evaluation. The other 506 processed reports comprise 500 Zoe reports outside the selected positional ranges, five unusable selected Zoe records and one unusable Maria record. The 2,000-row Zoe explanation artifact is a historical explanation-analysis surface, not the 1,894-report held-out evaluation population. Another supplied summary describes 2,493 reference-labeled and 45,545 largely unlabeled reports. That 2,493-report surface remains a separate external population until its manifest is admitted and reconciled.

**Endpoint crosswalk.** Core F1 is binary presence/absence after mapping levels 1–2 to absent and 3–4 to present. Whole-report accuracy requires all five decisions to match. Supplied exact-level "Certainty F1" remains distinct until its averaging formula is confirmed; it is not the native evaluator's exact four-level accuracy. On unlabeled reports, call rates and model agreement describe behavior, not correctness.

Generated explanations remain separate from structured predictions and deterministic metrics. They are candidate source phrases rather than clinical adjudications or access to internal reasoning. Patient-grouped inference remains unavailable because a validated report-to-patient map was not recovered.

#### H. Independent Mistral Reproduction

The independent reproduction is a separate evidence layer and does not replace submitted Mistral values. It used the author-confirmed intended configuration with exact model artifact SHA-256 `b85cdd596ddd76f3194047b9108a73c74d77ba04bef49255a50fc0cfbda83d32`, classification-prompt SHA-256 `52198221d8330e9857b51a7ad99b017aa18836e1718b08dd0ae355820f5a5e69`, explanation-prompt SHA-256 `09e7e46d13d9fd1f6ebd14e4caecf32766354ead048ef22aa834c5b6064cd05f`, classification-grammar SHA-256 `5237e13988062538cda9c21906f1f4e1fc8b99498e2462ea69fe24bface35016`, and explanation-grammar SHA-256 `718d3b0b`

`16499d04d97723893f5e1de67aa1f342ba8b455a293a3d93084cd315`.

The transport used raw completion prompts: task instructions and report text supplied directly, without a chat template marking user/model turns. Classification joined the prompt, two newline characters, and the report; explanation joined its prompt, the report, and the preceding classification in JSON (named fields with values). Reports were not silently shortened. Inputs exceeding the model's text-length limit would be reported as errors. Each report required two sequential calls.

Execution used an Apple M5 MacBook Pro with 24 GB unified memory, Python 3.11.15, `llama-cpp-python` 0.3.34, and Metal-backed llama.cpp. Zoe processing completed 2,000 reports in 51,246.6 seconds (25.63 seconds/report); Maria processing completed 500 reports in 12,617.2 seconds (25.22 seconds/report). Combined Mistral inference was approximately 17 hours 45 minutes. No observed prompt-plus-completion combination exceeded the 4,096-token context.

The run completed all 2,500 reports with zero missing or duplicate report identifiers, zero classification or explanation JSON/schema failures (wrong fields or structure), zero invalid four-level values, and no recorded text-length overflow or processing crash. Checking the meaning of those well-formed outputs nevertheless found activity/abnormality logical inconsistencies in 110/2,000 Zoe and 25/500 Maria outputs, explanation–decision mismatches in 1/10,000 Zoe and 2/2,500 Maria category decisions, and frequently formulaic explanations. These unfavorable observations describe the independent reproduction and are not used to alter the historical submitted results retrospectively.

### *I. MedGemma Evaluation with Raw-Completion Input*

Integration of MedGemma into the existing evaluation framework began with an independently specified MedGemma-27B text Q2_K run that sent the task and report as unmarked raw-completion text. It completed the same 1,395 Zoe and 499 Maria evaluation reports. All 1,994 processed records produced valid structured outputs with complete, unique keys: 100 transport-check records plus 1,894 evaluation records (1,395 Zoe and 499 Maria). The transport-check records are not included in the evaluation comparisons. This was a full evaluation, not only development or transport testing. The report-level comparison used exact same-case alignment, paired bootstrap intervals, McNemar tests, and Holm multiplicity adjustment. Across five labels, two cohorts, and comparisons with both submitted and reproduced Mistral, all 20 MedGemma-minus-Mistral 95% intervals for core-accuracy differences were below zero. Some false-negative-rate differences favored MedGemma, but these occurred with lower specificity and substantial overcalling. The lower agreement is specific to this raw-completion configuration and does not support a general conclusion about MedGemma. Patient-grouped inference remains unavailable without validated patient keys.

Table S5 reports the complete primary core-accuracy comparison rather than selecting a favorable category or cohort. It was generated deterministically from the validated aggregate analysis record.

### *J. MedGemma Evaluation with Model-Declared Input Serialization*

**What is compared here?**

Table S6 compares **MedGemma-27B Q2_K using its declared instruction-turn serialization with reproduced Mistral** on 1,894 matched evaluation reports. It tests portability of the report-to-result framework, not architectural superiority: the models differ in model family, parameter count and quantization. The preceding subsection reports MedGemma with unmarked raw-completion input. The bounded same-model check retained the MedGemma artifact, report keys, task, prompt, five categories, four-level labels, grammar, deterministic decoding, reference and endpoints; it changed only model-facing input serialization.

*Configured-system comparison.* The adapter placed the unchanged classification task and report inside the embedded user/model turn markers declared by the local MedGemma artifact. This was an ordinary local llama.cpp request, not a hosted-provider service. The output grammar remained active. A bounded eight-report replay manually assembled the same token sequence and reproduced the local chat-call labels and answer text on all eight cases; trimming whitespace alone did not do so. This supports a transport-level explanation—the model received its trained instruction-turn envelope—without identifying a single causal token or making a claim about hidden representations. A single configuration was selected before the 100-report development transport run by a structural rule that did not use reference metrics; the evaluation plan was then fixed before evaluation inference. The 100 development, 1,395 Zoe evaluation and 499 Maria evaluation records are pairwise disjoint by report key and normalized report text, and the 82-file transfer manifest was hash-verified. All 1,395 Zoe and 499 Maria reports were processed from the local model cache without report or prediction egress. Later development results did not select or alter this completed evaluation configuration.

Execution was entirely local on an Apple M5 with 24 GiB unified memory, using Python 3.11.15, `llama-cpp-python` 0.3.34 and Metal. The model was `medgemma-27b-text-it-Q2_K.gguf` from `unsloth/medgemma-27b-text-it-GGUF`, revision `334fbf6811c963d223f6ac107a459347353f068d`, SHA-256 `b137aac80f2bcb1c1ed35bfe13387bc496eb18898d5f46425687604f0f714481`. Model access was restricted to the local cache. All 1,894 classifications had valid schema and unique complete keys. Wall time was approximately eight hours, with mean classification inference time 15.14 seconds per report. The evaluation deliberately stopped after classification: it tests portability of the structured classification contract, not the historical pipeline's second-stage explanations. No cross-model explanation-quality or speed comparison is made. On the smaller Maria focal-epileptiform surface, the discordance pattern differed from Zoe: MedGemma alone matched the RA on five negative and three positive reports, versus three and one for Mistral; two false positives and two false negatives were shared between the models. These small groups do not support a stable mechanism.

TABLE S5: MedGemma-27B Q2_K evaluation using unmarked raw-completion input. Differences are MedGemma minus the named Mistral comparator on exactly aligned reports. CIs are 95% paired report-bootstrap intervals; $p_{\mathrm{Holm}}$ is the Holm-adjusted exact McNemar value.

| Cohort | Comparator | Label | MedGemma | Mistral | Difference (95% CI) | $p_{\mathrm{Holm}}$ |
|---|---|---|---|---|---|---|
| Zoe | Submitted Mistral | Abnormality | 0.798 | 0.961 | -0.163 (-0.186, -0.141) | $7.19 \times 10^{-44}$ |
| | | Focal Epi | 0.638 | 0.987 | -0.349 (-0.374, -0.323) | $3.78 \times 10^{-142}$ |
| | | Gen Epi | 0.784 | 0.968 | -0.184 (-0.207, -0.161) | $2.00 \times 10^{-50}$ |
| | | Focal Non-epi | 0.758 | 0.863 | -0.105 (-0.133, -0.077) | $7.85 \times 10^{-13}$ |
| | | Gen Non-epi | 0.781 | 0.899 | -0.118 (-0.141, -0.094) | $2.13 \times 10^{-20}$ |
| | Reproduced Mistral | Abnormality | 0.798 | 0.951 | -0.153 (-0.176, -0.129) | $4.85 \times 10^{-36}$ |
| | | Focal Epi | 0.638 | 0.987 | -0.349 (-0.374, -0.323) | $3.78 \times 10^{-142}$ |
| | | Gen Epi | 0.784 | 0.968 | -0.184 (-0.208, -0.161) | $4.28 \times 10^{-51}$ |
| | | Focal Non-epi | 0.758 | 0.872 | -0.114 (-0.143, -0.087) | $2.71 \times 10^{-15}$ |
| | | Gen Non-epi | 0.781 | 0.886 | -0.105 (-0.129, -0.080) | $7.68 \times 10^{-16}$ |
| Maria | Submitted Mistral | Abnormality | 0.788 | 0.916 | -0.128 (-0.172, -0.086) | $8.99 \times 10^{-8}$ |
| | | Focal Epi | 0.509 | 0.974 | -0.465 (-0.515, -0.419) | $1.81 \times 10^{-60}$ |
| | | Gen Epi | 0.758 | 0.986 | -0.228 (-0.267, -0.192) | $3.80 \times 10^{-31}$ |
| | | Focal Non-epi | 0.776 | 0.860 | -0.084 (-0.134, -0.038) | 0.002 |
| | | Gen Non-epi | 0.774 | 0.884 | -0.110 (-0.152, -0.070) | $2.97 \times 10^{-6}$ |
| | Reproduced Mistral | Abnormality | 0.788 | 0.904 | -0.116 (-0.160, -0.070) | $2.86 \times 10^{-6}$ |
| | | Focal Epi | 0.509 | 0.976 | -0.467 (-0.515, -0.421) | $1.02 \times 10^{-61}$ |
| | | Gen Epi | 0.758 | 0.988 | -0.230 (-0.269, -0.194) | $1.28 \times 10^{-32}$ |
| | | Focal Non-epi | 0.776 | 0.852 | -0.076 (-0.128, -0.026) | 0.007 |
| | | Gen Non-epi | 0.774 | 0.902 | -0.128 (-0.168, -0.088) | $5.39 \times 10^{-9}$ |

TABLE S6: MedGemma-27B Q2_K comparison using its declared instruction-turn serialization. Differences are MedGemma minus the named Mistral comparator on exactly aligned reports. Intervals are paired 95% report-bootstrap intervals; $p_{\mathrm{Holm}}$ is the Holm-adjusted exact report-level McNemar value.

| Cohort | Comparator | Label | MedGemma | Mistral | Difference (95% CI) | $p_{\mathrm{Holm}}$ |
|---|---|---|---|---|---|---|
| Zoe | Submitted Mistral | Abnormality | 0.945 | 0.961 | -0.016 (-0.029, -0.003) | 0.021 |
| | | Focal Epi | 0.974 | 0.987 | -0.013 (-0.020, -0.006) | 0.001 |
| | | Gen Epi | 0.989 | 0.968 | 0.021 (0.011, 0.029) | $2.69 \times 10^{-5}$ |
| | | Focal Non-epi | 0.913 | 0.863 | 0.050 (0.029, 0.070) | $1.50 \times 10^{-5}$ |
| | | Gen Non-epi | 0.941 | 0.899 | 0.042 (0.026, 0.057) | $1.79 \times 10^{-6}$ |
| | Reproduced Mistral | Abnormality | 0.945 | 0.951 | -0.006 (-0.020, 0.009) | 0.501 |
| | | Focal Epi | 0.974 | 0.987 | -0.013 (-0.020, -0.006) | 0.001 |
| | | Gen Epi | 0.989 | 0.968 | 0.020 (0.011, 0.029) | $6.04 \times 10^{-5}$ |
| | | Focal Non-epi | 0.913 | 0.872 | 0.041 (0.021, 0.061) | $2.50 \times 10^{-4}$ |
| | | Gen Non-epi | 0.941 | 0.886 | 0.054 (0.038, 0.071) | $1.67 \times 10^{-9}$ |
| Maria | Submitted Mistral | Abnormality | 0.950 | 0.916 | 0.034 (0.004, 0.064) | 0.120 |
| | | Focal Epi | 0.984 | 0.974 | 0.010 (-0.004, 0.024) | 0.680 |
| | | Gen Epi | 0.986 | 0.986 | 0.000 (-0.008, 0.008) | 1.000 |
| | | Focal Non-epi | 0.954 | 0.860 | 0.094 (0.060, 0.126) | $3.05 \times 10^{-7}$ |
| | | Gen Non-epi | 0.946 | 0.884 | 0.062 (0.036, 0.088) | $1.56 \times 10^{-5}$ |
| | Reproduced Mistral | Abnormality | 0.950 | 0.904 | 0.046 (0.016, 0.076) | 0.013 |
| | | Focal Epi | 0.984 | 0.976 | 0.008 (-0.006, 0.022) | 0.775 |
| | | Gen Epi | 0.986 | 0.988 | -0.002 (-0.010, 0.004) | 1.000 |
| | | Focal Non-epi | 0.954 | 0.852 | 0.102 (0.068, 0.134) | $4.16 \times 10^{-8}$ |
| | | Gen Non-epi | 0.946 | 0.902 | 0.044 (0.022, 0.066) | 0.001 |

Table S6 retains all 20 core comparisons with submitted and reproduced Mistral, always using the same reports for both models. Three different questions underlie the statistics:

- **How large is the difference, and how much might it vary?** We repeatedly resample reports with replacement, keeping both models' predictions and the reference together for each selected report. These 2,000 paired report-bootstrap replicates (seed 20260718) give the displayed 95% intervals without rerunning a model. Each interval concerns one difference, not all comparisons jointly.
- **Does one model match the reference more often?** We use an exact matched-pairs test for binary outcomes, conventionally called **McNemar's test** [3]. The test compares cases where only MedGemma matches with cases where only the comparator matches. Cases where both match or neither matches do not favor either model. In formal terms, this tests marginal homogeneity of the paired correctness proportions. Our exact two-sided calculation is the equivalent binomial test on the two directions among these discordant pairs. The null hypothesis is that the directions occur equally often; a small $p$-value indicates evidence against that balance, not the probability that a model is wrong. Here, "correct" means matching the reference annotation, not independently verified clinical truth.
- **How do we limit chance findings when asking several questions?** We use Holm's step-down multiple-testing adjustment (also called the Holm–Bonferroni method) [4]. It makes the $p$-value threshold stricter across ten tests per cohort and comparator: five labels times

binary and exact four-level agreement. Binary agreement checks present versus absent (levels 1–2 versus 3–4); exact four-level agreement requires the same level from 1 to 4. For example, Zoe native MedGemma versus submitted Mistral forms one ten-test group. Under the tests' assumptions, Holm limits the chance of any false statistical finding in that group to 5%: mistakenly declaring a difference when the no-difference hypothesis is true. Adjustment was not across all study comparisons: it provides no single study-wide error guarantee.

The full source-hashed aggregate ledger—a results register linked to exact source-file fingerprints—retains all 90 core, ordinal and false-negative-rate effects, including the separate SA comparisons. The displayed intervals are not Holm-adjusted; interval direction and adjusted $p$-values therefore answer different questions. Patient-grouped inference remains unavailable: the analysis cannot yet account for several reports belonging to the same patient. Neither pairing models on reports nor Holm adjustment resolves that dependence.

Core pointwise intervals—intervals for individual comparisons—were above zero for 12 comparisons, below zero for three and included zero for five. Above zero favors MedGemma, below zero favors the named Mistral comparator, and an interval including zero is also compatible with no difference. These are not independent wins. For example, the positive Maria abnormality interval versus submitted Mistral did not pass the Holm-adjusted threshold ($p = 0.120$): the single-comparison interval favored MedGemma, but the test did not meet the stricter multiple-question threshold. All favorable, null and adverse categories are retained.

**Reading the focal-epileptiform trade-off.** For Zoe focal epileptiform activity, MedGemma identified all 53 RA-positive reports, whereas reproduced Mistral missed two. Among 1,342 RA-negative reports, positive calls that disagreed with the RA increased from 16 (1.2%) to 36 (2.7%). Precision consequently decreased from 76.1% to 59.6%, and Core Agreement from 98.7% to 97.4%. Here, "false positive" means that the model assigned a positive label where the RA assigned absence; it is not an independently adjudicated clinical error. For research cohort construction, this configuration would retrieve the two additional RA-positive reports but send twenty additional RA-negative reports to review. Whether that trade-off has clinical significance requires qualified case review.

Other trade-offs were also retained. Zoe overall-abnormality false positives increased from 14 to 53 while misses decreased from 55 to 24. On Maria, focal non-epileptiform misses decreased from 57 to three while false positives increased from 17 to 20. These comparisons concern agreement with the study reference, not clinical correctness.

*Why can the same task give different answers?:* **Input formatting and output constraints do different jobs.** The model receives tokens, not just the conceptual question. MedGemma's declared instruction-turn format marks the user turn, its end, and where the model's answer should begin. These markers are part of the input format described in Google's Gemma documentation; they are not merely visual punctuation. The decoding grammar separately restricts the answer's structure and label choices. It cannot supply missing input-turn structure or guarantee semantic correctness.

**The local connector, in plain language.** The raw-completion call supplies the task followed by the report. The declared-format adapter trims outer whitespace and encloses that same content in a user turn, followed by the start of a model turn. The installed library then invokes its ordinary completion engine with those tokens and the same output grammar. Model-declared serialization therefore does not mean unconstrained output, an interactive conversation, or a remote service. The user/model turn structure can also be assembled explicitly and submitted through the completion interface; beginning-of-sequence tokens must not be duplicated.

**A small controlled replay, not another evaluation.** Eight fixed, spaced development reports were replayed in five formats (40 calls), keeping the model, task, grammar and sampling fixed:

- **Declared-format call versus hand-assembled completion:** identical input tokens and answer text on all eight reports.
- **Restoring historical stopping behavior:** no answer changed from the assembled declared-format input.
- **Trimming outer whitespace alone:** labels changed on two reports, but still differed from declared-format labels on three reports.

All 40 outputs were valid; the original raw-completion and declared-format classifications each reproduced their saved labels on all eight reports. No reference labels or evaluation reports were used. This identifies a reproducible connector adjustment while retaining constrained outputs. It does not isolate every turn marker's effect, explain the model's internal reasoning, establish a new accuracy estimate, or show that all classification errors are fixed. Full prompts and same-report outputs remain in the controlled diagnostic record.

#### K. Small Mistral Interface Sensitivity

**Same Mistral model; two input formats**

This section compares **Mistral-7B with itself**, using raw-completion input versus its own declared instruction format on 100 development reports. It asks whether the same adapter principle also helps Mistral. It is not a new full-cohort Mistral–MedGemma comparison.

Only the input serialization was changed in a capped 100-report development check; the Mistral artifact, task prompt, grammar and sampling were retained. Declared-format minus raw-completion Core Agreement changes were $-1$, $0$, $-2$, $-3$ and $+3$ percentage points for abnormality, focal epileptiform, generalized epileptiform, focal non-epileptiform and generalized non-epileptiform activity, respectively. All five paired core intervals included zero and all adjusted core $p$-values were 1.0. This is no consistent observed gain, not equivalence; no full-cohort declared-format Mistral rerun followed.

**Plain-language interpretation.** The changes were small and went in both directions. This check gives no reason to assume that Mistral needs the same repair as MedGemma. It does not establish that the two Mistral interfaces are interchangeable on every report, or explain the model-internal cause of their differing sensitivity.

TABLE S7: **Effect of removing the endpoint-confidence instruction from the Mistral-7B prompt.** One-factor ablation specified in a committed protocol before execution on the fixed 100-report Zoe development set (500 category decisions); all other pipeline elements held fixed. The columns are not the complete first and final thesis prompts. Not a restoration of the historical "up to 14%" claim.

| Measure (500 decisions) | Present | Removed | Difference |
|---|---|---|---|
| Endpoint labels 1 or 4 | 418/500 (83.6%) | 396/500 (79.2%) | −4.4 points |
| Binary Core Agreement | 475/500 (95.0%) | 473/500 (94.6%) | −0.4 points |
| Exact four-level agreement | 411/500 (82.2%) | 388/500 (77.6%) | −4.6 points |

*What did the endpoint-confidence instruction change?:* The recovered method-development record identifies one component contrast as "V2: V1 + Confidence Guideline." Table S7 does not compare the complete first and final prompts reproduced in the thesis's Prompt Tuning chapter [5], which differ in several components. Instead, it isolates the named confidence-guidance component. The guidance-present condition is the saved first-100 subset of the independently reproduced final Mistral raw-completion run. The guidance-removed condition is a new run in which only the following two sentences were deleted: "Err on the side of confident decisions. Use 1 or 4 whenever possible. Only use 2 or 3 if there is strong, unavoidable ambiguity. Choose 2 or 3 sparingly, only when absolutely necessary." The model artifact, raw-completion interface, all remaining prompt text, grammar, deterministic decoding, report order and RA labels remained fixed. All 100 first Zoe development reports were used; neither evaluation cohort nor the explanation stage was accessed.

The configurations made the same binary decision on 486/500 category decisions but the same four-level assignment on 459/500. For focal epileptiform activity, all 100 binary decisions were unchanged, while 13 assignments moved from level 1 to 2 and one from level 4 to 3 after guidance removal. Cross-field violation counts were unchanged. Thus the instruction primarily shaped expressed certainty within the absent/present halves of the scale, rather than repairing semantic consistency or consistently changing the binary finding decision.

**Boundary.** This is a development sensitivity, not held-out evidence, probability calibration or clinical validation. It neither reconstructs nor replaces the removed historical "up to 14%" claim. It shows only that endpoint guidance shaped ordinal output.

### *L. Limitations*

Reports came from one regional health authority and two report-author surfaces; Maria is not an external institution. Analyses are report-level because a validated report-to-patient map was unavailable. The RA is a study reference, not diagnostic ground truth, and rare positive categories limit precision. Exact four-level agreement is not probability calibration. Historical prompt stopping, baseline fold assignments, and fitted models were not fully retained. The Mistral and MedGemma configurations differ in model family, size, quantization, and required input serialization, so their comparison does not isolate a base-model effect. No prospective workflow or clinical-benefit evaluation was performed.
